## Supplementary material for "ChatGPT- versus human-generated answers to frequently asked questions about diabetes: a Turing test-inspired survey among employees of a Danish diabetes center": R Markdown with code and results

Adam Hulman

2023-02-10

##### Context

This document includes R code used for data processing and analysis, and results reported in in the study by Hulman et al. entitled *ChatGPT-generated versus human expert-written answers to frequently asked questions about diabetes: a Turing test-inspired e-survey among all employees of a Danish diabetes center.* The study protocol had been published on Figshare (link) before data collection began.

##### R session info

```
library(table1)
```

```
##
## Attaching package: 'table1'
```

```
## The following objects are masked from 'package:base':
##
##     units, units<-
```

```
library(flextable)
library(miceadds)
```

```
## Loading required package: mice
```

```
##
## Attaching package: 'mice'
```

```
## The following object is masked from 'package:stats':
##
##     filter
```

```
## The following objects are masked from 'package:base':
##
##     cbind, rbind
```

```
## * miceadds 3.16-18 (2023-01-06 10:54:00)
```

```
library(Epi)
```

```
##
## Attaching package: 'Epi'
```

```
## The following object is masked from 'package:flextable':
##
##     before
```

```
library(lme4)
```

```
## Loading required package: Matrix
```

```
##
## Attaching package: 'lme4'
```

```
## The following object is masked from 'package:Epi':
##
##     factorize
```

```
sessionInfo()
```

```
## R version 4.2.2 (2022-10-31 ucrt)
## Platform: x86_64-w64-mingw32/x64 (64-bit)
## Running under: Windows 10 x64 (build 19042)
##
## Matrix products: default
```

```
##
## locale:
## [1] LC_COLLATE=English_United States.utf8
## [2] LC_CTYPE=English_United States.utf8
## [3] LC_MONETARY=English_United States.utf8
## [4] LC_NUMERIC=C
## [5] LC_TIME=English_United States.utf8
##
## attached base packages:
## [1] stats     graphics  grDevices utils     datasets  methods   base
##
## other attached packages:
## [1] lme4_1.1-31      Matrix_1.5-1     Epi_2.47         miceadds_3.16-18
## [5] mice_3.15.0      flextable_0.8.5  table1_1.4.2
##
## loaded via a namespace (and not attached):
##  [1] tidyr_1.2.1         jsonlite_1.8.3      splines_4.2.2
##  [4] Formula_1.2-4       shiny_1.7.4         askpass_1.1
##  [7] yaml_2.3.6          gdtools_0.3.0       numDeriv_2016.8-1.1
## [10] pillar_1.8.1        backports_1.4.1     lattice_0.20-45
## [13] glue_1.6.2          uuid_1.1-0          digest_0.6.30
## [16] promises_1.2.0.1    minqa_1.2.5         cmprsk_2.2-11
## [19] htmltools_0.5.4     httpuv_1.6.8        plyr_1.8.8
## [22] gfonts_0.2.0        pkgconfig_2.0.3     httpcode_0.3.0
## [25] broom_1.0.2         purrr_1.0.1         xtable_1.8-4
## [28] later_1.3.0         officer_0.5.2       tibble_3.1.8
## [31] openssl_2.0.4       mgcv_1.8-41         generics_0.1.3
## [34] ellipsis_0.3.2      etm_1.1.1           cachem_1.0.6
## [37] cli_3.4.1           survival_3.4-0      magrittr_2.0.3
## [40] crayon_1.5.2        mime_0.12           memoise_2.0.1
## [43] evaluate_0.18       fansi_1.0.3         nlme_3.1-160
## [46] MASS_7.3-58.1       xml2_1.3.3          tools_4.2.2
## [49] data.table_1.14.4   mitools_2.4         lifecycle_1.0.3
## [52] stringr_1.4.1       zip_2.2.2           compiler_4.2.2
## [55] systemfonts_1.0.4   rlang_1.0.6         grid_4.2.2
## [58] nloptr_2.0.3        rstudioapi_0.14     base64enc_0.1-3
## [61] rmarkdown_2.18      boot_1.3-28         DBI_1.1.3
## [64] curl_4.3.3          R6_2.5.1            zoo_1.8-11
## [67] knitr_1.41          dplyr_1.0.10        fastmap_1.1.0
## [70] utf8_1.2.2          stringi_1.7.8       parallel_4.2.2
## [73] crul_1.3            Rcpp_1.0.9          vctrs_0.5.0
## [76] tidyselect_1.2.0    xfun_0.34
```

#### Data processing

```
dataset_all <- read.csv(file_path,
                    sep = ',',
                    dec = '.')

dataset_complete <- subset(dataset_all, stato_4==1 | stato_3==1)

dataset_complete$id <-1:nrow(dataset_complete)
```

```
variables <- c('id', 'age', 'sex', 'contact', 'heard', 'used',
               paste0('q_',1:10))

dataset_wide <- dataset_complete[, variables]

dataset_wide$id <- factor(dataset_wide$id)

dataset_wide$age30 <- factor(dataset_wide$age,
                             levels = c(2,1,3,4),
                             labels = c('30-39',
                                        'under 30',
                                        '40-49',
                                        'over 50'))

dataset_wide$age50 <- factor(dataset_wide$age,
                             levels = c(4,1,2,3),
                             labels = c('over 50',
                                        'under 30',
                                        '30-39',
                                        '40-49'))

dataset_wide$age <- factor(dataset_wide$age,
                           levels = 1:4,
                           labels = c('under 30',
                                      '30-39',
                                      '40-49',
                                      'over 50'))

dataset_wide$sex <- factor(dataset_wide$sex,
                           levels = c(0, 1),
                           labels = c('female',
                                      'male'))

dataset_wide$contact <- factor(dataset_wide$contact,
                               levels = c(0, 1),
                               labels = c('no',
                                          'yes'))

dataset_wide$heard <- factor(dataset_wide$heard,
                             levels = c(0, 1),
                             labels = c('no',
                                        'yes'))

dataset_wide$used[is.na(dataset_wide$used)] <- 0
dataset_wide$used <- factor(dataset_wide$used,
                            levels = c(0, 1),
                            labels = c('no',
                                       'yes'))

dataset_long <- reshape(dataset_wide,
                        direction = 'long',
```

```
                        varying = paste0('q_',1:10),
                        sep = '_',
                        timevar = 'question',
                        v.names = "correct")

dataset_long$question <- factor(dataset_long$question)
```

#### Table 1

```
t1flex(table1(~ age + contact + heard + used | sex, data=dataset_wide))
```

```
## Warning: fonts used in `flextable` are ignored because the `pdflatex` engine
## is used and not `xelatex` or `lualatex`. You can avoid this warning by using
## the `set_flextable_defaults(fonts_ignore=TRUE)` command or use a compatible
## engine by defining `latex_engine: xelatex` in the YAML header of the R Markdown
## document.
```

| | female (N=129) | male (N=52) | Overall (N=183) |
|---|---|---|---|
| **age** | | | |
| under 30 | 11 (8.5%) | 5 (9.6%) | 18 (9.8%) |
| 30-39 | 47 (36.4%) | 23 (44.2%) | 70 (38.3%) |
| 40-49 | 44 (34.1%) | 12 (23.1%) | 56 (30.6%) |
| over 50 | 27 (20.9%) | 12 (23.1%) | 39 (21.3%) |
| **contact** | | | |
| no | 61 (47.3%) | 15 (28.8%) | 76 (41.5%) |
| yes | 68 (52.7%) | 37 (71.2%) | 107 (58.5%) |
| **heard** | | | |
| no | 57 (44.2%) | 7 (13.5%) | 66 (36.1%) |
| yes | 72 (55.8%) | 45 (86.5%) | 117 (63.9%) |
| **used** | | | |
| no | 119 (92.2%) | 27 (51.9%) | 148 (80.9%) |
| yes | 10 (7.8%) | 25 (48.1%) | 35 (19.1%) |

#### Main analysis

```
coef_to_prob <- function(x) 1/(1+exp(-x)) # inverse logit

model <- miceadds::glm.cluster(data=dataset_long,
                               formula = correct ~ 1,
                               cluster = "id",
                               family = "binomial")
```

```
## Loading required namespace: sandwich
```

```
summary(model)
```

```
##               Estimate Std. Error  z value     Pr(>|z|)
## (Intercept) 0.3857739 0.05260449 7.333478 2.242554e-13
```

```
est_ci <- ci.lin(model)[c(1,5,6)]
est_ci_prob <- coef_to_prob(est_ci)
round(est_ci_prob, 4) # overall
```

```
## [1] 0.5953 0.5702 0.6198
```

#### Secondary analyses (univariable models)

##### Age

```
model_age <- miceadds::glm.cluster(data=dataset_long,
                                   formula = correct ~ age,
                                   cluster = "id",
                                   family = "binomial")

summary(model_age)
```

```
##                Estimate Std. Error     z value   Pr(>|z|)
## (Intercept)  0.37001836  0.1747845  2.11699709 0.03426009
## age30-39    -0.05872642  0.1926096 -0.30489879 0.76044324
## age40-49     0.01090564  0.2030370  0.05371258 0.95716416
## ageover 50   0.17078810  0.2028536  0.84192783 0.39982835
```

```
est_ci_age <- ci.lin(model_age, ctr.mat = rbind(c(1,0,0,0), # under 30
                                                c(1,1,0,0), # 30-39
                                                c(1,0,1,0), # 40-49
                                                c(1,0,0,1)) # over 50
                     )[,c(1,5,6)]

est_ci_prob_age <- coef_to_prob(est_ci_age)
row.names(est_ci_prob_age) <- levels(dataset_long$age)
round(est_ci_prob_age, 4)
```

```
##          Estimate   2.5%  97.5%
## under 30   0.5915 0.5069 0.6710
## 30-39      0.5772 0.5381 0.6154
## 40-49      0.5941 0.5445 0.6419
## over 50    0.6320 0.5840 0.6776
```

##### Sex

```
model_sex <- miceadds::glm.cluster(data=dataset_long,
                                   formula = correct ~ sex,
                                   cluster = "id",
                                   family = "binomial")

summary(model_sex)
```

```
##              Estimate Std. Error  z value     Pr(>|z|)
## (Intercept) 0.3135842   0.060014 5.225184 1.739824e-07
## sexmale     0.2422046   0.121767 1.989083 4.669208e-02
```

```
est_ci_sex <- ci.lin(model_sex, ctr.mat = rbind(c(1,0),             # female
                                                c(1,1))             # male
                     )[,c(1,5,6)]

est_ci_prob_sex <- coef_to_prob(est_ci_sex)
row.names(est_ci_prob_sex) <- levels(dataset_long$sex)
round(est_ci_prob_sex, 5)
```

```
##        Estimate    2.5%   97.5%
## female  0.57776 0.54883 0.60616
## male    0.63548 0.58616 0.68210
```

**Patient contact**

```
model_contact <- miceadds::glm.cluster(data=dataset_long,
                                       formula = correct ~ contact,
                                       cluster = "id",
                                       family = "binomial")

summary(model_contact)
```

```
##              Estimate Std. Error  z value     Pr(>|z|)
## (Intercept) 0.2923880 0.07570807 3.862045 0.0001124419
## contactyes  0.1618673 0.10440212 1.550422 0.1210403343
```

```
est_ci_contact <- ci.lin(model_contact, ctr.mat = rbind(c(1,0),       # no
                                                        c(1,1))       # yes
                         )[,c(1,5,6)]

est_ci_prob_contact <- coef_to_prob(est_ci_contact)
row.names(est_ci_prob_contact) <- levels(dataset_long$contact)
round(est_ci_prob_contact, 5)
```

```
##     Estimate    2.5%   97.5%
## no   0.57258 0.53594 0.60844
## yes  0.61165 0.57770 0.64455
```

**ChatGPT use**

```
model_used <- miceadds::glm.cluster(data=dataset_long,
                                    formula = correct ~ used,
                                    cluster = "id",
                                    family = "binomial")

summary(model_used)
```

```
##              Estimate Std. Error  z value     Pr(>|z|)
## (Intercept) 0.3080660 0.05634155 5.467830 4.555795e-08
## usedyes     0.4204831 0.13946114 3.015056 2.569319e-03
```

```
ci.exp(model_used)
```

```
##             exp(Est.)     2.5%   97.5%
## (Intercept)  1.360791 1.218522 1.51967
## usedyes      1.522697 1.158521 2.00135
```

```
est_ci_used <- ci.lin(model_used, ctr.mat = rbind(c(1,0),             # no
                                                  c(1,1))             # yes
                      )[,c(1,5,6)]

est_ci_prob_used <- coef_to_prob(est_ci_used)
row.names(est_ci_prob_used) <- levels(dataset_long$used)
round(est_ci_prob_used, 5)
```

```
##     Estimate    2.5%   97.5%
## no   0.57641 0.54925 0.60312
## yes  0.67449 0.61740 0.72683
```

### Figure 1

```
pch_par <- 19
cex_par <- 1.3
cex_text <- 0.8
cex_text2 <- 0.7

plot_figure <- function(){
  plot(9,9,
     main=NULL,
     xlab="%",
     ylab="",
     bty="n",
     xaxt="n",
     yaxt="n",
     ylim=c(0,16),
     xlim=c(0.4,1))
```

```r
  polygon(c(0.40,0.40,0.55,0.55), c(17.65,-1,-1,17.65),
          col =rgb(0, 153, 180,maxColorValue = 255),border='transparent',
          density = 5, angle = 45)
  abline(v = seq(0.4,1,0.1), col = "lightgray", lwd = 0.2, lty = 1)
  axis(1,at=seq(0.4,1,0.01), labels = F, tck = -0.02)
  axis(1,at=c(0.55,seq(0.4,1,0.1)), labels = c(55,seq(40,100,10)))

  abline(v = 0.55, col = rgb(0, 153, 180, maxColorValue = 255), lwd = 2, lty = 1)
  text(0.475,0, 'non-inferiority zone', cex = cex_text,
       col = rgb(0, 153, 180, maxColorValue = 255))

  segments(est_ci_prob[2],1,est_ci_prob[3],1)
  points(est_ci_prob[1],  1, pch=pch_par, cex = cex_par)
  text(0.91,1,'overall', adj = 0, cex = cex_text, font = 2)

  segments(est_ci_prob_used[,2],4:3,est_ci_prob_used[,3],4:3)
  points(est_ci_prob_used[,1],4:3, pch=pch_par, cex = cex_par)
  text(0.91,4,'ever used', adj = 0, cex = cex_text)
  text(0.91,3,'ChatGPT', adj = 0, cex = cex_text)
  text(0.81,4,'no', adj = 0, cex = cex_text2)
  text(0.81,3,'yes', adj = 0, cex = cex_text2)

  segments(est_ci_prob_contact[,2],7:6,est_ci_prob_contact[,3],7:6)
  points(est_ci_prob_contact[,1],7:6, pch=pch_par, cex = cex_par)
  text(0.91,7,'patient', adj = 0, cex = cex_text)
  text(0.91,6,'contact', adj = 0, cex = cex_text)
  text(0.81,7,'no', adj = 0, cex = cex_text2)
  text(0.81,6,'yes', adj = 0, cex = cex_text2)

  segments(est_ci_prob_sex[,2],10:9,est_ci_prob_sex[,3],10:9)
  points(est_ci_prob_sex[,1],10:9, pch=pch_par, cex = cex_par)
  text(0.91,9.5,'sex', adj = 0, cex = cex_text)
  text(0.81,10,'female', adj = 0, cex = cex_text2)
  text(0.81,9,'male', adj = 0, cex = cex_text2)

  segments(est_ci_prob_age[,2],15:12,est_ci_prob_age[,3],15:12)
  points(est_ci_prob_age[,1],15:12, pch=pch_par, cex = cex_par)
  text(0.91,13.5,'age', adj = 0, cex = cex_text)
  text(0.81,15,'< 30', adj = 0, cex = cex_text2)
  text(0.81,14,'30-39', adj = 0, cex = cex_text2)
  text(0.81,13,'40-49', adj = 0, cex = cex_text2)
  text(0.81,12,'> 50', adj = 0, cex = cex_text2)
}

plot_figure()
```

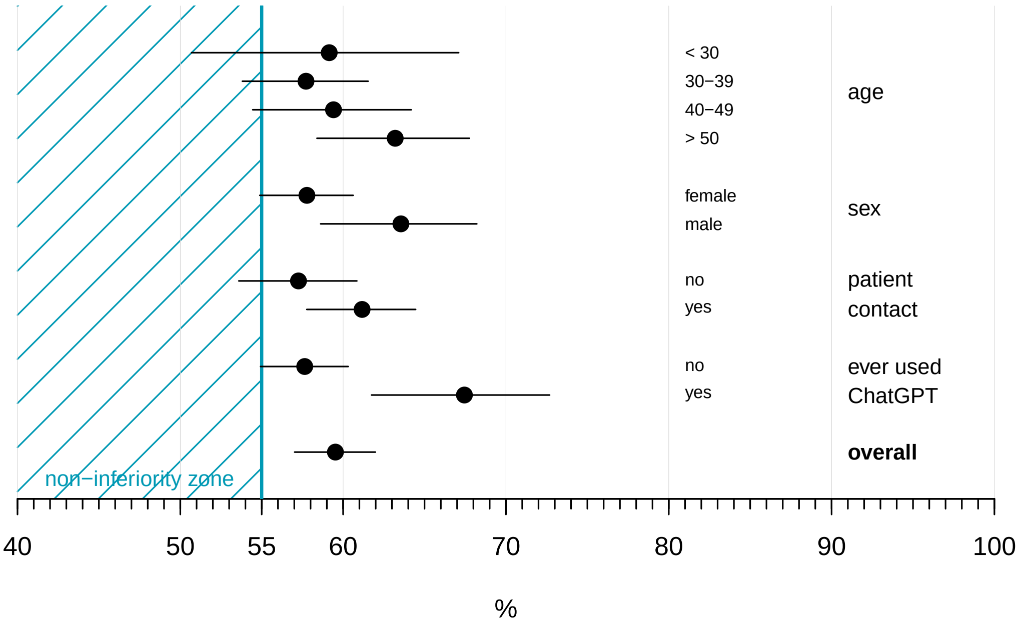

#### Exploratory analyses

##### Multivariable model

```
model_comb <- miceadds::glm.cluster(data=dataset_long,
                                    formula = correct ~ age + sex + contact + used,
                                    cluster = "id",
                                    family = "binomial")

summary(model_comb)
```

```
##                 Estimate Std. Error    z value    Pr(>|z|)
## (Intercept)  0.15961295  0.1912616  0.8345271 0.403984035
## age30-39    -0.04785869  0.2047092 -0.2337887 0.815149021
## age40-49     0.06846088  0.2136870  0.3203792 0.748680894
## ageover 50   0.21203493  0.2124911  0.9978531 0.318350604
## sexmale      0.05908650  0.1230938  0.4800120 0.631218855
## contactyes   0.14546042  0.1041176  1.3970778 0.162390184
## usedyes      0.41025487  0.1469434  2.7919249 0.005239551
```

```
#odds ratios
ci.exp(model_comb)
```

```
##             exp(Est.)      2.5%    97.5%
## (Intercept) 1.1730568 0.8063376 1.706558
```

```
## age30-39    0.9532685 0.6382142 1.423849
## age40-49    1.0708587 0.7044358 1.627882
## ageover 50  1.2361911 0.8151036 1.874815
## sexmale     1.0608670 0.8334572 1.350326
## contactyes  1.1565720 0.9430779 1.418397
## usedyes     1.5072019 1.1300379 2.010249
```

```
# model with age 30-39 as reference category
model_comb2 <- miceadds::glm.cluster(data=dataset_long,
                                    formula = correct ~ age30 + sex + contact + used,
                                    cluster = "id",
                                    family = "binomial")

summary(model_comb2)
```

```
##                   Estimate Std. Error   z value    Pr(>|z|)
## (Intercept)     0.11175426  0.1041537 1.0729748 0.283282409
## age30under 30 0.04785869  0.2047092 0.2337887 0.815149021
## age3040-49     0.11631957  0.1274247 0.9128494 0.361321734
## age30over 50  0.25989361  0.1259599 2.0633048 0.039083676
## sexmale        0.05908650  0.1230938 0.4800120 0.631218855
## contactyes     0.14546042  0.1041176 1.3970778 0.162390184
## usedyes        0.41025487  0.1469434 2.7919249 0.005239551
```

```
# odds ratios
ci.exp(model_comb2)
```

```
##                 exp(Est.)      2.5%    97.5%
## (Intercept)      1.118238 0.9117557 1.371482
## age30under 30  1.049022 0.7023216 1.566872
## age3040-49       1.123355 0.8750902 1.442052
## age30over 50    1.296792 1.0131019 1.659922
## sexmale          1.060867 0.8334572 1.350326
## contactyes       1.156572 0.9430779 1.418397
## usedyes          1.507202 1.1300379 2.010249
```

##### Between-person variation

```
model_ranef <- glmer(correct ~ 1 + (1 | id),
                   data = dataset_long,
                   family = binomial)

summary(model_ranef)
```

```
## Generalized linear mixed model fit by maximum likelihood (Laplace
##   Approximation) [glmerMod]
##  Family: binomial  ( logit )
## Formula: correct ~ 1 + (1 | id)
##    Data: dataset_long
##
```

```
##      AIC      BIC   logLik deviance df.resid
##   2395.9   2406.9  -1196.0   2391.9     1772
##
## Scaled residuals:
##     Min      1Q  Median      3Q     Max
## -1.3443 -1.1791  0.7688  0.8208  0.9053
##
## Random effects:
##  Groups Name        Variance Std.Dev.
##  id     (Intercept) 0.07766  0.2787
## Number of obs: 1774, groups:  id, 183
##
## Fixed effects:
##             Estimate Std. Error z value Pr(>|z|)
## (Intercept)  0.39284    0.05332   7.368 1.73e-13 ***
## ---
## Signif. codes:  0 '***' 0.001 '**' 0.01 '*' 0.05 '.' 0.1 ' ' 1
```

```
fixed_int <- as.numeric(fixef(model_ranef))
ranef_int <- as.data.frame(ranef(model_ranef))$condval

qqnorm(fixed_int + ranef_int)
```

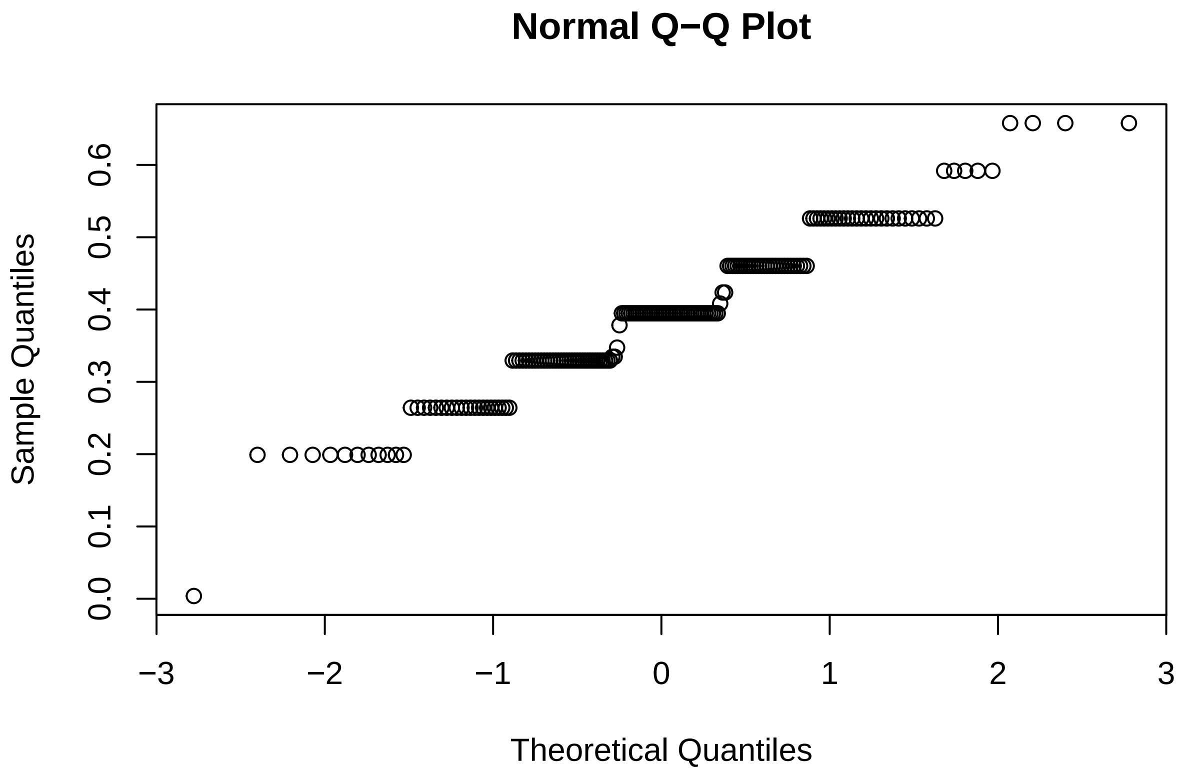

```r
# 95% prediction interval
coef_to_prob(c(0.39284 - 1.96*0.2787, 0.39284 + 1.96*0.2787))
```

```
## [1] 0.4617220 0.7189162
```

##### Results by questions

```r
q_text <- c('Hvor meget frugt må jeg spise, når jeg har diabetes?',
            'Skal jeg justere min insulinbehandling, når jeg er syg med feber?',
            'Hvordan opbevarer jeg insulin på en lang rejse?',
            'Skal jeg være bekymret for mine fødder, når jeg har diabetes?',
            'Hvorfor er mine blodsukre hoeje?',
            'Hvordan påvirker motion blodsukkeret når man har type 1 diabetes?',
            'Kan light-sodavand få mit blodsukker til at stige og påvirke min diabetes?',
            'Kan diabetes påvirke sexlivet?',
            'Hvordan påvirker forskellige former for træning typisk blodsukkeret
             hos personer med type 1 diabetes?',
            'Hvad er graviditetsdiabetes?')

model_question <- miceadds::glm.cluster(data=dataset_long,
                                    formula = correct ~ question,
                                    cluster = "id",
                                    family = "binomial")

summary(model_question)
```

```
##                Estimate Std. Error     z value     Pr(>|z|)
## (Intercept)  0.66865616  0.1566127  4.26948984 1.959206e-05
## question2    0.17864170  0.2091189  0.85425894 3.929615e-01
## question3   -1.15867866  0.2113794 -5.48151230 4.217054e-08
## question4   -0.75962794  0.2161389 -3.51453525 4.405242e-04
## question5   -0.10904037  0.2240569 -0.48666379 6.264966e-01
## question6    0.01598033  0.2214841  0.07215115 9.424816e-01
## question7   -0.32438661  0.2136271 -1.51847152 1.288956e-01
## question8    0.39987867  0.2184577  1.83046255 6.718080e-02
## question9   -0.78241505  0.2271502 -3.44448318 5.721522e-04
## question10  -0.13351323  0.2298596 -0.58084682 5.613437e-01
```

```r
Q_mat <- cbind(1,rbind(0,diag(9)))

est_ci_question <- ci.lin(model_question, ctr.mat = Q_mat)[,c(1,5,6)]

est_ci_prob_question <- coef_to_prob(est_ci_question)
row.names(est_ci_prob_question) <- levels(dataset_long$question)
prob_table_question <- round(est_ci_prob_question, 4)
# probabilities by question
# first column indicates the position of the question in the survey
prob_table_question[order(prob_table_question[,1]),]
```

```
##    Estimate   2.5%  97.5%
```

```
## 3    0.3799 0.3116 0.4533
## 9    0.4716 0.3988 0.5456
## 4    0.4773 0.4043 0.5512
## 7    0.5852 0.5109 0.6559
## 10   0.6307 0.5568 0.6989
## 5    0.6364 0.5626 0.7042
## 1    0.6612 0.5895 0.7262
## 6    0.6648 0.5917 0.7308
## 2    0.7000 0.6289 0.7626
## 8    0.7443 0.6746 0.8035
```

```
# questions in increasing order by probability
print(q_text[order(prob_table_question[,1])],row.names = F)
```

```
##  [1] "Hvordan opbevarer jeg insulin på en lang rejse?"
##  [2] "Hvordan påvirker forskellige former for træning typisk blodsukkeret \n             hos persone
##  [3] "Skal jeg være bekymret for mine fødder, når jeg har diabetes?"
##  [4] "Kan light-sodavand få mit blodsukker til at stige og påvirke min diabetes?"
##  [5] "Hvad er graviditetsdiabetes?"
##  [6] "Hvorfor er mine blodsukre hoeje?"
##  [7] "Hvor meget frugt må jeg spise, når jeg har diabetes?"
##  [8] "Hvordan påvirker motion blodsukkeret når man har type 1 diabetes?"
##  [9] "Skal jeg justere min insulinbehandling, når jeg er syg med feber?"
## [10] "Kan diabetes påvirke sexlivet?"
```
